# Service User and Healthcare-worker Perspectives on Substance Use Disorder Treatment Journeys, Relapse, and Recovery in Sub-Saharan Africa: Evidence from Uganda

**DOI:** 10.64898/2025.12.08.25341874

**Authors:** Claire Biribawa, Wouter Vanderplasschen, Joan Nankya Mutyoba, Byamah Brian Mutamba, Kenneth Kalani, Daniel Oyet, David Kalema, Nazarius Mbona Tumwesigye, Vicki Simpson

**Author notes:** Corresponding Author: Claire Biribawa.

## Abstract

**Introduction:** Substance Use Disorders (SUDs) pose significant public health challenges in sub-Saharan Africa. However, there is limited evidence on how service users (SUs) and front-line health care workers (HCWs) experience SUD treatment and recovery within resource constrained settings. This study explored the perspectives of SUs and HCWs to understand treatment trajectories and to identify multilevel barriers and facilitators influencing recovery outcomes.

**Methods:** We conducted a qualitative study at two public treatment facilities in Uganda; the Alcohol and Drug Unit at Butabika National Referral Mental Hospital and the Mental Health Unit at Gulu Regional Referral Hospital. Using purposive sampling, we interviewed 10 HCWs directly involved in SUD care (five per site) and randomly selected 43 SUs from a parent cohort of 439 adults treated for SUDs at the same sites. We transcribed all audio recordings verbatim and translated them into English where necessary. We coded SUs and HCWs data separately and then applied integrative mapping to generate cross-cutting meta-themes.

**Results:** We identified four interrelated meta-themes that described how individual, social, community and health system factors influenced treatment journey and recovery outcomes. Both SUs and HCWs highlighted resource constrained service environments characterized by shortages of specialist staff, medications and secure infrastructure, which compelled clinicians to improvise and resulted in fragmented care. Human resource gaps contributed to rushed assessments, minimal psychosocial engagement, and limited follow-up. Treatment pathways were described as heavily biomedical, with services centred on detoxification and management of acute withdrawal, while psychosocial needs, and contextual stressors insufficiently addressed. Outside the treatment facility, individual, social and community barriers like stigma, strained family relationships, peer pressure, poverty, alcohol-dependent livelihoods, and untreated mental or physical comorbidities made recovery difficult, as SUs returned to the same environments that precipitated their substance use. Despite these challenges, participants identified opportunities to strengthen recovery pathways, including integration of family and spiritual support, involvement of “expert-client” models, anti-stigma interventions, and task-sharing with community health workers.

**Conclusion:** Sustained recovery from SUDs in Uganda is constrained by fragmented clinical services, systemic resource shortages, stigma, and unsupportive social contexts. Improving recovery therefore demands a multilevel response including strengthening psychosocial support, integrating family and spiritual support, peer role models, anti-stigma initiatives and task-sharing with community health workers to strengthen recovery pathways and sustain long-term treatment outcomes.

## Introduction

Substance use disorders (SUDs) constitute one of the fastest growing yet persistently neglected, drivers of global morbidity and mortality, contributing substantially to Disability-Adjusted Life Years (DALYs)(1–6). The World Drug Report 2024 estimates that in 2020, about 292 million people worldwide used illicit drugs which is a 20% increase over the previous decade while approximately 64 million met criteria for a drug use disorder (DUD), representing a 3% rise over the past five years (7). The rising incidence of DUDs is primarily driven by population growth and socio-economic factors, with opioids being a major contributor to mortality and DALYs (8,9). Alcohol Use Disorders (AUDs), however, still account for the largest proportion of the global burden of SUDs. According to the Global status report on alcohol and health and treatment of substance use disorders, an estimated 400 million people, about 7% of the world’s population aged 15 years and older, live with AUDs(10). These trends are unfolding against a backdrop of widening treatment gaps, constrained health care budgets, and shifting drug markets dominated by cheap, highly potent synthetics.

The burden of harmful substance use is rising in sub-Saharan Africa (SSA). The World Health Organization’s regional office for Africa estimates that alcohol alone accounts for 3.3 million deaths annually, and at least 15 million live with DUDs in the region(11). Alcohol consumption rates in SSA are among the highest globally, with an estimated per capita consumption of 9.4 liters of pure alcohol per year(12,13). Prevalence of AUDs varies widely across countries ranging from 0.1% to 33.2% in SSA(14,15) though the true burden is likely underestimated due to underreporting and a high rate of undocumented alcohol consumption(14). Alongside heavy alcohol use, the region is now contending with rapidly growing illicit drug use including opioids such as tramadol, methamphetamine, and home-brewed concoctions such as “kush”, “nyaope”, “gaddafi”, among others which combine pharmaceuticals, solvents, and plant materials with often lethal consequences (16–20).

Access to formal treatment services in SSA remains limited. Most formal treatment services are concentrated in urban areas, leaving rural populations underserved(21). Care is often fragmented, ranging from limited residential and inpatient facilities to, emerging primary care based on brief interventions, along with widespread traditional and spiritual healing practices that serve as first-line care options(22,23).

Uganda exemplifies the challenges embedded in Africa’s changing substance-use landscape. The Country ranks among the highest globally for per capita alcohol consumption, at 12.2 liters per person annually(24), with approximately 10% of the adult population estimated to have an AUD(25,26). Uganda is also seeing a surge in use of other drugs including cannabis, codeine, and amphetamine-type stimulants(27–30). The situation is further compounded by a healthcare system which is ill-equipped to address the needs of individuals with SUDs. The treatment landscape is characterized by limited residential facilities concentrated in urban areas, gaps in service delivery capacity, and continued reliance on traditional healing practices(31–33). Limited access to quality healthcare, coupled with stigma and cultural perceptions of addiction further constrain effectiveness of interventions(34).

Despite the recognized importance of expanding SUD treatment in SSA, significant research gaps remain. Most studies have examined either patient experiences or health system capacity in isolation, limiting understanding of the dynamic interactions between service delivery and service utilization. Few empirical studies in SSA have explored how service users themselves understand and mobilize such available resources or how front-line providers navigate their absence. This dual approach is essential for understanding mismatches between provider capacity and attitudes on one side and patient needs and expectations on the other, which often undermine treatment engagement and outcomes(35). To address this gap, we conducted a qualitative study that integrated the perspectives of health-care workers (HCWs) and service users (SUs) into a single analytic frame to map the full SUD-care trajectory from admission through community re-entry and to identify multilevel barriers and facilitators that shape treatment and recovery outcomes.

## Materials and Methods

### Ethics Statement

Ethical approval was obtained from Makerere University School of Public Health Research Ethics Committee (MakSPH-REC) as part of the AQUALEFF study (Advancing Quality and Effectiveness of Treatment of Substance Use Disorders in Uganda), prior to data collection. The study was subsequently registered with the Uganda National Council for Science and Technology in accordance with national research guidelines under Registration Number HS6699ES. Additionally, administrative clearance to conduct research at the treatment facilities was obtained.

All participants provided written informed consent prior to data collection. Participation was voluntary, and confidentiality and anonymity were assured throughout the study. Data, including transcripts and audio recordings were securely stored in encrypted digital folders, accessible only to the core research team, to ensure data integrity and confidentiality. Ethical guidelines were strictly adhered to during all stages of the research.

### Study Design

This study utilized a qualitative research design employing in-depth interviews with SUs seeking treatment and key-informant interviews with HCWs directly involved in the treatment and care of individuals with SUDs. The study adopted a constructivist paradigm, drawing on constructivist grounded theory to guide both data collection and analysis(36–39). The study is reported in accordance with the 32-item Consolidated Criteria for Reporting Qualitative Research (40).

### Study Setting

The research study was conducted at two public hospitals in Uganda: Butabika National Referral Mental Health Hospital (BNRMH) in Kampala, and the mental-health unit of Gulu Regional Referral Hospital (GRRH) in northern Uganda. BNRMH is the national tertiary centre for mental health and substance-use care. Within BNRMH, the Alcohol and Drug Unit (ADU) functions as Uganda’s only government - run rehabilitation centre for substance-use disorders, offering a full continuum of care: medically supervised detoxification; structured inpatient rehabilitation - with a bed capacity of approximately 80 patients delivered on a standard 2–4-week protocol with step-down planning; and post-discharge outpatient follow-up via scheduled clinics and community linkage. The service pathway begins with detoxification (medical stabilization of withdrawal symptoms under specialist supervision), followed by a rehabilitation phase involving inpatient care where psychosocial counselling, relapse-prevention planning, group therapy, family involvement and vocational re-integration sessions are provided. Once patients transition from the inpatient phase, the unit offers outpatient follow-up services, comprising scheduled clinics for monitoring, individual and group counselling, peer-support groups and referral back into community-based care or outreach services. The ADU also houses Uganda’s first Medically Assisted Therapy clinic for opioid use disorder which serves hundreds of clients annually through combined harm-reduction and recovery-oriented care. The staffing mix at BNRMH combines psychiatrists, psychiatric clinical officers, psychiatric nurses, clinical psychologists, counsellors, social workers and peer-support mentors, together forming a multidisciplinary team responsible for assessment, care-planning, treatment implementation and discharge planning.

GRRH serves as the primary mental health regional referral center for districts in the Acholi sub-region (including the districts of Amuru, Gulu, Kitgum, Lamwo and Pader). GRRH Mental Health Unit is integrated within the general hospital and provides inpatient (approximately 20 beds) and outpatient psychiatric services, managed by psychiatrists and psychiatric clinical officers with linkages to district and community services.

Together, these two settings provided a valuable contrast between national and regional service delivery models. This distinction is important for interpreting differences in service-pathways and experiences between the two sites. All interviews were carried out face-to-face in private consultation rooms at the two public facilities.

### Participants, Sampling methods and Sample size

We included two categories of participants: (i) patients seeking treatment for SUDs referred to as service users (SUs), and (ii) HCWs directly involved in the treatment and care of individuals with SUDs. SUs were drawn from a parent cohort of 439 adults treated for SUDs at the two public health facilities in Uganda. Individuals were eligible for this qualitative study if they were aged 18 years or older, had a clinician-confirmed SUD diagnosis according to DSM-5 criteria (41), and had engaged with treatment services as part of their care. Individuals presenting with acute psychosis or severe cognitive impairment that precluded informed consent were excluded from participation but were offered appropriate referrals for further care.

To minimize investigator bias while preserving heterogeneity within the cohort, we used a computer-generated random number to select 45 SUs. Of these, 43 SUs consented to participate, while 2 declined due to time constraints. The sample size was determined pragmatically considering available time and resources however this number was sufficient to capture a wide range of perspectives and enable in-depth exploration of treatment experiences.

Among the 43 SUs, 31 were male and 12 were female, with the mean age of 35 years (range 19– 55 years). The most reported primary substances of use were alcohol (n = 17), cannabis (n = 14), illegal opioids (n =7), legal opioids such as tramadol and pethidine (n=4), and sedative-hypnotics (n=1). Polysubstance use was reported by 20 participants (46.5%). For the service providers, we employed a purposive sampling strategy to recruit 10 HCWs directly involved in the care of individuals with SUD at the study sites. They included clinical psychologists (n=4), psychiatric nurses (n=2), counselors (n=2), and community health workers (CHWs) (n=2) with firsthand experience in delivering SUD-related services. Their median age was 34 years, and most were male (70%).

### Data Collection

We collected data through in-depth individual interviews with SUs and HCWs from 2^nd^ September 2023 – 25 March 2025 and from 17 June 2025 – 28 August 2025. The first author, together with trained research assistants, conducted all interviews in private rooms at the participating facilities to ensure confidentiality and minimize disruptions.

Interviews were guided by semi-structured interview guides that were tailored separately for SUs and HCWs to capture their distinct experiences and perspectives on SUD treatment, relapse, and recovery. An interview guide was developed based on the study objectives, literature review, and expert input. It included open-ended prompts covering themes on treatment experiences, recovery journeys, pathways to care, healthcare system barriers, and recommendations for improving treatment outcomes. While the guide ensured consistency, interviews remained flexible to accommodate emergent topics and participant narratives. The full interview guides for HCWs and SUs are provided in Appendix 1.

Interviews were conducted in English, Luganda, Acholi, or Swahili, depending on participants’ language preferences. No interpreters were required, as the research assistants were fluent in the respective local languages. Each interview lasted approximately 45 minutes to 60 minutes and was audio-recorded with participants’ consent. Interviewers also took brief field notes to document contextual observations and non-verbal cues that complemented the transcribed data. All audio recordings were transcribed verbatim and, where necessary, translated into English to preserve meaning and expression of participants’ accounts. Transcripts were reviewed for completeness and accuracy before analysis.

### Data Analysis

We conducted the analysis within a constructivist paradigm, guided by the principles of constructivist grounded theory, and applied Braun and Clarke’s six-step framework for thematic analysis (42). This approach enabled us to identify patterns and meanings across participant narratives and to explore how individual and contextual factors influenced treatment journeys and outcomes.

In the first step, two independent researchers with backgrounds in public health (CB) and psychology (VS) read and re-read all transcripts to gain a comprehensive understanding of the content and record preliminary impressions. We analyzed interviews from SUs and HCWs separately in the initial stages to preserve context-specific meanings.

We developed initial codes manually through close reading of the transcripts, highlighting meaningful text segments and documenting emerging concepts and analytic reflections. The researchers met regularly to compare and refine the coding framework, reviewing codes against extracts to ensure coherence and consistency.

We then clustered related codes into sub-themes, which represented closely connected ideas or processes within each participant group’s accounts. After developing sub-themes separately for SUs and HCWs, we conducted a comparative analysis to examine how these sub-themes aligned, overlapped, or contrasted across the two datasets. Through iterative team discussions and integrative mapping, we synthesized these findings into a set of cross-cutting meta-themes that captured shared experiences as well as group-specific insights on treatment, relapse, and recovery. We assigned clear definitions and labels to each sub-theme and meta-theme to ensure they accurately reflected the underlying data and participant narratives.

## Results

We provide detailed findings from 53 in-depth interviews conducted with 43 SUs receiving treatment for SUDs and 10 HCWs providing direct care in two public treatment facilities in Uganda. Table 1 provides an overview of four interrelated meta-themes and sub-themes reflecting participants’ experiences and perspectives across the continuum of SUD treatment, relapse, and recovery (Table 1).

**Table 1:**
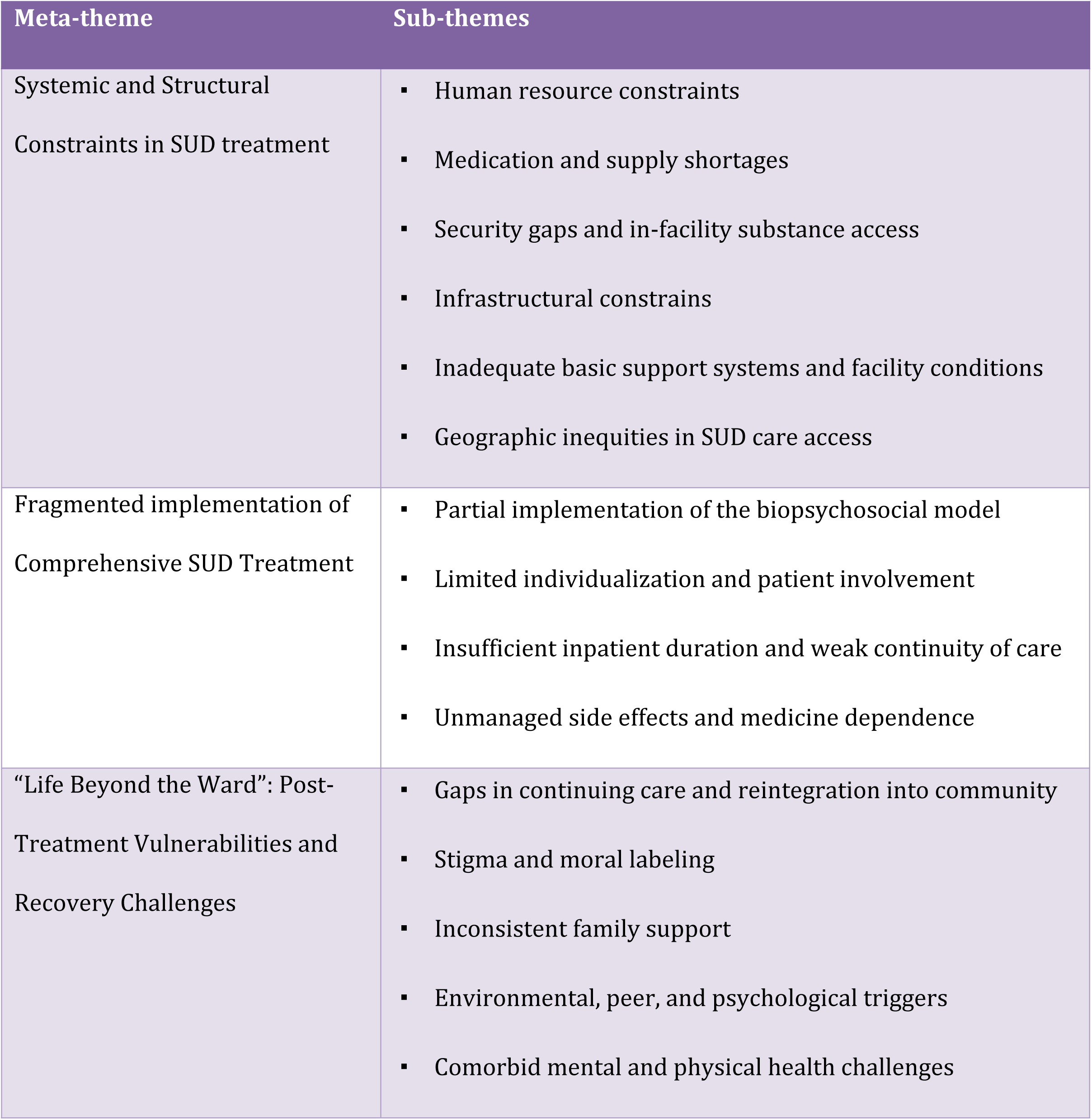
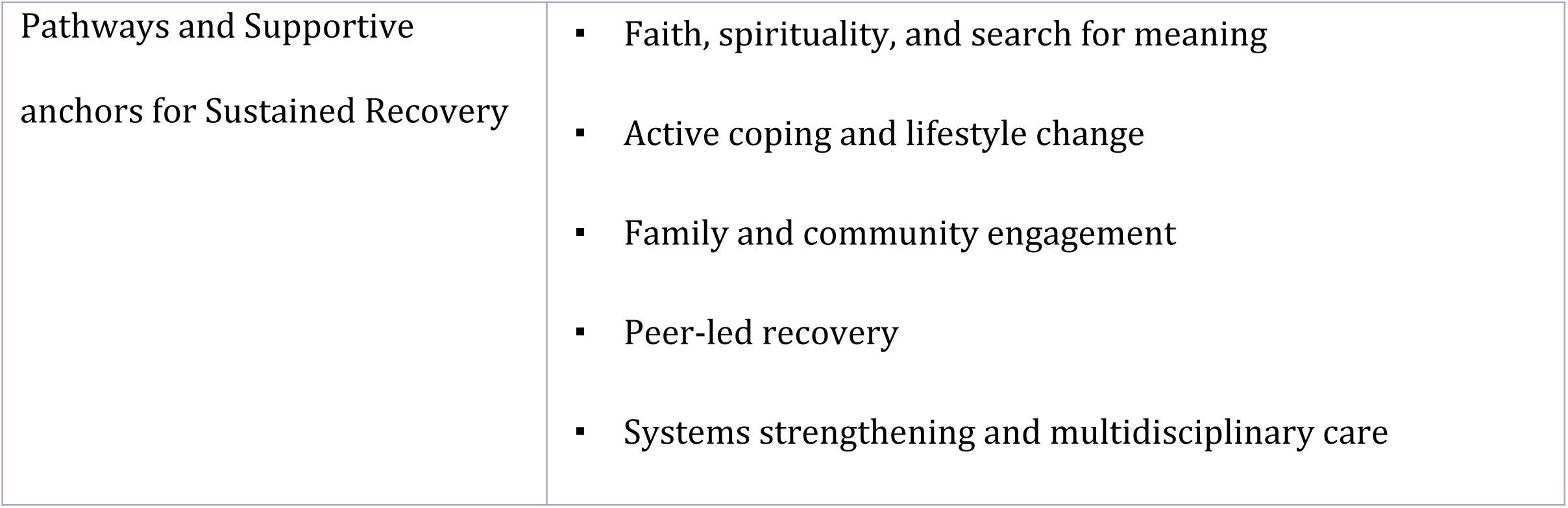
Overview of meta-themes and sub-themes derived from service users and healthcare workers on substance use disorder treatment and recovery in Uganda.

### Meta-theme 1: Systemic and structural constraints in SUD treatment

This meta-theme captured the overarching narrative of an overstretched and under-resourced system struggling to provide consistent and specialized SUD care. It highlighted six sub-themes: human resource constraints, medication and supply shortages, security and in-facility substance access, infrastructural challenges, inadequate basic support systems and facility conditions and geographical inequities in SUD access to care.

### Sub-theme 1.1: Human resource constraints

At both study sites, participants described severe staff shortages and high patient-to-provider ratios, leading to work overload, burnout, and compromised quality in delivery of services. Facilities operated with minimal specialist staff such as counselors, psychologists, and occupational therapists. HCWs reported taking on multiple roles, often beyond their training or capacity.

> *“Human Resource is one of major challenges we face here at the unit, for example, there is no attached counsellor or psychologist at the unit. So as the clinical team we try to fill up these gaps but haphazardly.” (HCW #3)*

These staffing pressures, coupled with stockouts of essential medicines, constrained clinical effectiveness, deepened providers’ sense of helplessness, and contributed to moral distress among those who recognized what holistic, patient-centered treatment should entail but were unable to deliver it. The recurrence of “revolving-door” admissions where patients relapsed shortly after discharge reinforced feelings of futility, as staff viewed these outcomes as consequences of systemic limitations rather than individual failings.

> *“Sometimes you get tired, you feel like you’re doing the same thing over and over again; you treat someone, they go out, and a few months later they come back… and yet there are more patients out there, and bed space is not increasing.” (HCW#4)*.

Participants also highlighted poor remuneration, limited incentives, and slow career progression as additional demotivators.

> *“Someone can take even 10 years to be promoted… and yet you have the qualifications. So, there’s the way it demoralizes even the people who are working.” (HCW#6)*

### Subtheme 1.2: Medication and supply shortages

Both HCWs and SUs at both facilities reported persistent stockouts of medications such naltrexone which disrupted continuity of care and forced clinicians to improvise with non-pharmacological interventions such as counselling, behavioral therapy or symptomatic management which can be less effective.

> *“When the patient cannot afford to buy some of these medications… we divert to psychological measures such as distraction, exercise, sleep medication.” (HCW#5)*

SUs also described being asked to purchase medications externally when hospital supplies ran out, often at prohibitive costs. These shortages deepened inequities, leaving patients who could not afford without adequate treatment options which reinforced their perceptions that care was inconsistent and unreliable.

> *“They tell you to go buy this medicine because it is not there at the hospital, you reach the pharmacy, and it is very expensive.” (SU#11)*

### Sub-theme 1.3: Security gaps and in-facility substance access

HCWs at BNRMH reported inadequate security screening procedures, which facilitated the smuggling of psychoactive substances into inpatient wards by patients and visitors.

> *“We don’t have a good security… drugs sneak in…so you are trying to treat the patient… you find that the patient is high… when you do a drug screen you find that he has used drugs while still on the ward.”* (HCW#4)

At GRRH, the mental health unit’s open layout with no perimeter wall allowed patients to move out freely to access substances or engage with community members using substances.

> *“Our unit is an open space, and patients come in and go out whenever they want. Some sneak in substances and so we are detoxing, but the person is taking more substances. I remember a patient whom the peers smuggled weed for him (HCW#2)*

At both facilities such lapses undermined treatment integrity, created mistrust between staff and patients, and demoralized HCWs who felt that efforts to support recovery were being undone within the facility itself.

### Sub-theme 1.4: Infrastructural Constraints

Across both facilities, participants described overcrowding, inadequate bed space, and lack of private areas for counseling. In both facilities, patient numbers frequently exceeded available capacity, resulting in the placement of some individuals in acute psychiatric wards that were not designed for addiction rehabilitation. These and other structural deficiencies undermined comprehensive care.

> *“Some of our patients… are willing to leave alcohol… they are calm… they are not supposed to on the acute ward because it is for psychotic cases.” (HCW#8)*
>
> *“When facilities become overwhelmed by bed capacity, patients are discharged at the first sign of slight improvement, even if they aren’t clinically ready.” (HCW#2)*

### Sub-theme 1.5: Deficits in essential patient support and facility living conditions

Both SUs and HCWs from GRRH emphasized that poor living conditions and unmet basic needs particularly the lack of food directly undermined patient retention and recovery. HCWs highlighted that some inpatients were discharged prematurely or absconded because they could not get meals during their stay. In the absence of institutional food provision, patients often left the ward to seek food, returning re-intoxicated or failing to return at all, thereby interrupting detoxification and increasing the risk of relapse.

> *“Right now, conditions in these facilities are really challenging, for example, patients aren’t even provided with food so they are discharged prematurely because they cannot afford to be at the facility or they just run away.” (HCW#3)*

In addition to food insecurity, participants described poor sanitation that reduced patient comfort.

> *“The toilets keep blocking and they become very dirty, so it is very uncomfortable to be admitted here….” (SU#14)*

### Sub-theme 1.6 Geographic inequities in SUD care access

Both HCWs and SUs highlighted that publicly funded SUD services were highly centralized restricting access for patients from rural and distant districts. HCWs mentioned that although BNRMH was intended to serve the entire Country, there was no structured referral systems with limited linkage with lower-level facilities. Similarly, at GRRH, HCWs that SUD care was concentrated at regional level, while lower health system levels (Health Centre II–IV) remained under-resourced contributing to late presentation and relapse. SUs also echoed these concerns, emphasizing the need for accessible, decentralized services.

> *“The health system levels from general hospital right to Health Centre II are not helping. People by-pass all those levels and come to the regional referral and yet the distance and transport costs are very high, they come from far villages.” (HCW #1)*
>
> *“The center should be in all government hospitals because many people aren’t aware that help can be got for addiction problems.” (SU #14)*

### Meta-theme 2: Fragmented implementation of comprehensive SUD treatment

This meta-theme highlights the fragmented and inconsistently implemented nature of substance use disorder (SUD) care across treatment facilities. Despite Uganda’s treatment framework being conceptually grounded in a biopsychosocial model, its operationalization was partial, with biomedical interventions disproportionately dominating service delivery. Participants described minimal patient engagement in treatment planning, brief inpatient admissions that constrained recovery consolidation, and inadequate management of medication-related side effects. Collectively, these accounts underscore a discontinuous, non-individualized, and insufficiently holistic approach to SUD care.

### Sub-theme 2.1: Partial implementation of the Biopsychosocial model

While HCWs at BNRMH and GRRH acknowledged a biopsychosocial approach in principle, both HCWs and SUs agreed that treatment delivery was dominated by biomedical and pharmacological interventions. The psychological and social aspects of care such as counseling, occupational therapy, and family engagement were inconsistently implemented due to systemic resource constraints like under staffing and other logistical challenges.

> *“Some of the Health Workers are treating the symptoms…they ignore counseling, spiritual healing and yet these are equally important… (HCW#3)*
>
> *“When I came here, I was not encouraged to stop using substances, I did not get any counselling…. I thought I was just forced to stop taking marijuana.” (SU#9)*

### Sub-theme 2.2: Limited individualization and patient involvement in treatment

Accounts from both HCWs and SUs highlighted a lack of individualized care and patient participation in the treatment process. Participants described SUD care as largely standardized, with limited collaboration and adaptation to individual needs. High caseloads limited time for comprehensive assessments and developing personalized treatment plans.

> *“…we do not do thorough assessments to critically understand the categories of our patients. Some of our patients are brought to the unit tied but we fail to classify them and treat them in the one size fits all way.” (HCW#3)*

Likewise, SUs reported minimal involvement in decision-making, with some entering treatment under family pressure or receiving medication without adequate explanation.

> *“I was forced to take medication which I do not agree with and instead of it helping me, it just hindered me because I do not agree with taking pills when I am not sick.” (SU#6)*

### Sub-theme 2.3: Insufficient in-patient duration and transition in care

Across facilities, HCWs reported that inpatient treatment durations were shorter than they considered adequate for sustained recovery. At BNRMH, they noted stays averaged at 2–2.5 months but were often truncated due to high patient turnover and limited bed capacity. At GRRH, HCWs described typical admissions of about two weeks, acknowledging that such brief stays were insufficient for meaningful behavioral and psychosocial stabilization.

> *“… we tend to have a huge number of patients waiting to be admitted to the ADU and sometimes when you feel the patient has been able to get this and that, we just continue with him as an outpatient to create some space.” (HCW#9)*
>
> *“The time provided to the patients is very low… two weeks is not sufficient to treat these addictions. We just wait for them to come back.” (HCW#1)*

### Sub-theme 2.4: Unmanaged side-effects and medicine dependence

Service users reported experiencing distressing side-effects such as sedation, drowsiness, sexual dysfunction, physical discomfort, and disturbed sleep often without adequate explanation or management by clinicians. Some questioned whether treatment itself was harmful and expressed anxiety about developing dependence on prescribed medications. These concerns eroded trust in the therapeutic process and discouraged sustained engagement with treatment.

> *“I feel addicted to these tablets, because medication is something that replaced drinking. You feel like you are a slave to this medication.” (SU#12)*
>
> *“…… the hallucinations continued coming and I decided to take alcohol to help me deal with them which spoilt my sobriety, and I was taken back ….” (SU#4)*

### Meta-theme 3: “Life beyond the ward”: post-treatment vulnerabilities and recovery challenges

This meta-theme captured how recovery did not occur in a vacuum but extended beyond the hospital setting, unfolding within challenging social and structural environments. Reintegration often meant returning to the same environments and networks that had enabled substance use, posing major challenges to sustained recovery. Participants described gaps in continuing care, stigma and moral labeling, inconsistent family involvement, and peer, environmental, and psychological triggers increasing relapse risk.

### Sub-theme 3.1: Gaps in continuing care and community reintegration

Across facilities, participants described major gaps in post-discharge follow-up and community reintegration. At BNRMH, HCWs described a clear intention to continue care after discharge through reviews, family sessions, peer/community arms, and even home visits but were rarely implemented due to resource constraints.

> *“In our treatment plan, we’re even supposed to do home visits… but with limited staff and many patients… we are unable to do this.” (HCW#4)*

Further they reported that community and peer structures such as the Uganda Harm Reduction Network and AA/NA-type groups reached only a small proportion of discharged service users, mainly around Kampala. Consequently, most received minimal or no continuing care, contributing to relapse and repeat admissions that clogged an already strained inpatient service. Similar challenges were reported at GRRH, where HCWs cited absence of relapse prevention strategies, family engagement, or the establishment of community resources to provide ongoing support leaving recovery largely dependent on individual effort rather than coordinated support.

> *“There is no pro-active follow-up of the patients in the communities to understand how they are managing. We just sit back and wait for the patients, hoping they will come back for review.” (HCW #2)*

### Sub-theme 3.2: Stigma and moral labelling

Both HCWs and SUs reported how moralistic perceptions, stigmatizing language, and labeling undermined SUD recovery. Stigma arose from various sources including HCWs, families, and communities. SUD was often viewed as a moral failing or sign of madness, leaving participants feeling rejected, misunderstood, and ashamed.

> *“These client’s express willingness to quit, but upon returning to their communities, they face rejection…they are still looked at as addicts and the rest of the community does not want to associate.” (HCW#8)*

Terms such as “abuser,” sometimes used by HCWs, reinforced negative stereotypes and caused emotional distress while internalized stigma led to shame and social withdrawal. HCWs emphasized the importance of adopting more non-stigmatizing terminology.

> *“When I went back home, people still called me names, mad, addict… because they thought I was still using.” (SU#17)*

### Sub-theme 3.3: Inconsistent family support in SUD care

Reports from participants indicated that family involvement varied widely. Supportive families played a crucial role in motivating recovery. Some SUs reported that their families offered emotional, logistical, or financial support which motivated recovery.

> *“what I can say is my family members, they supported me a lot, when I relapsed, they rush me to hospital…when I don’t have money, they buy for me medicine.” (SU#13)*

However, supportive families were the exception. By the time patients reached treatment, many families were emotionally and financially exhausted, with some withdrawing entirely.

> *“The family members sometimes greatly contribute to the relapse of the patient, by the time the person comes at the facility they are treated as rejects and the relatives have been pushed to the limits. It is rare that the family comes to support.” (HCW #2)*

Even well-intentioned families sometimes contributed to relapse. Poor communication, unresolved trauma, emotionally charged encounters, and family stressors such as marital breakdowns destabilized recovery. Despite their importance, structured family involvement in SUD treatment was largely absent.

> *“When you have people who are generally judgmental, then I tell myself maybe let me have a beer first. Then you reach home and your wife starts shouting… Now you’ve gone back to drinking and others are just there discouraging you.” (SU#4)*

### Sub-theme 3.4: Environmental and peer-driven triggers

Both HCWs and SUs indicated that recovery was challenged by peer pressure and environments saturated with alcohol and drug use. Respondents consistently reported that the moment they left the ward they were “thrown back into a sea of substances.” Some participants explained that local brewing is a source of livelihood and making recovery hard.

> *“I have a patient who is always coming back to the facility with alcohol induced psychosis…her employment is brewing local alcohol… She usually tells me that you cannot sell what you have not tasted.” (HCW#9)*

Many SUs described being drawn back into familiar social networks where substance use remained normalized. SUs’ companions dismissed HCW advice and offered substances as gestures of friendship, reactivating cravings and undermining progress made.

> *“My friends refuted HCWs’ statements against my drinking abstinence…saying they are just foolish and that alcohol does nothing to a person…” (SU#24)*

### Sub-theme 3.5: Social and psychological triggers

Service users reported that emotional distress, social isolation, and daily stressors were major triggers for relapse. Many lacked supportive relationships and spent long periods alone, which intensified cravings and depression. Feelings of anger, loneliness, and frustration exacerbated by stigma, weak family support, unstable employment, and financial hardship created persistent psychological strain. For some, incarceration further disrupted community reintegration, and so substance use was a quick escape route from despair.

> *“My life was all about, marijuana, cannabis, khat, and alcohol, it reached a time when I had come back from prison, with loads of thoughts and felt alone. I attempted suicide but the knife felt painful… so I went into the bush with poison thinking that I would have my last marijuana then drink the poison, but after smoking the marijuana, I felt life became meaningful again.” (SU#32)*

Participants linked stress and economic insecurity to feelings of hopelessness and idleness, which in turn reinforced cravings and relapse.

### Sub-theme 3.6: Comorbid mental and physical health challenges

Both HCWs and SUs reported that co-occurring mental health comorbidities such as depression, anxiety, bipolar disorder, among others complicated SUDs treatment and undermined recovery. HCWs also noted that unaddressed medical comorbidities further complicated care, as SUs reported living with chronic medical illnesses that increased everyday stress and triggered cravings. SUs described concerning challenges related to polysubstance use, with some receiving treatment for only one substance despite using several concurrently.

> *“…Even when you run my blood sugar levels right now, they are high, and it depresses me… then I get episodes of hypoglycemia, I start sweating and then people start laughing at me that that is alcohol –waragi! Stupid, I am dying here, and you are busy telling me that.” (SU#4)*

### Meta-theme 4: Pathways and supportive anchors for sustained recovery

Despite these barriers participants identified a spectrum of intrinsic, social, spiritual, and community anchors that could be mobilized to sustain recovery. These were analyzed into four sub-themes which included intrinsic and spiritual motivation, active coping and lifestyle change, family and community collaboration, and systems strengthening.

### Sub-theme 4.1: Faith, spirituality and search for meaning in recovery

Participants frequently described that faith, spiritual belief, prayer, and religious community engagement served as powerful internal motivators for change and sustained recovery.

> *“I listen to Christian radio called ‘Favour FM’, it really encourages me not to take alcohol. They talk about experiences of people, and this helps me not to be like those people that ended up in bad situations.” (SU#30)*
>
> *“…I decided to turn to religion to help me deal with the condition. In Islam, we pray 5 times in a day, and it helps me to remain sober.” (SU#32)*

Both Christian and Muslim participants described religion as a stabilizing force that helped them resist peer pressure, reframe their purpose, and provided a sense of belonging and frameworks for daily discipline, self-reflection, and community accountability.

> *“Currently I have been managing the church public address system, and this keeps me busy, and I don’t have a lot of free time to go use the drugs. In case I have any challenges, I share them with my pastor, and I get words of encouragement and advice.” (SU#7)*

Respondents advocated for the inclusion of faith leaders, Village Health Teams, and other local community groups in the formal recovery programs.

> *“….. what I think is just one, what you people (medics) do is not bad its good, but we should allow a religious person to be a part of the team, even if you people do the counselling…, sometimes you may be far from the person and yet the person is going through some hardship. It may be hard for the person to come to hospital to seek counseling, it would be better if a religious leader is nearby because these days there are many churches around so during such times the religious leader can be providing some help while the person waits for the appointment date to return to the hospital. Am recommending this because I have seen the benefits from my personal life…” (SU#7)*

Parental duty also emerged as a powerful internal driver. Respondents reported that thinking first about basic needs for their children helped redirect money away from alcohol.

> *“These days, when I get some money, I first think about getting food for my children before anything else.” (SU#30)*

### Sub-theme 4.2: Active coping and lifestyle change

Service users emphasized the benefit of adopting new, structured routines that replaced substance-related behaviors. Keeping physically busy, engaging in meaningful work, and staying focused on long-term responsibilities were repeatedly highlighted as self-directed tools for maintaining sobriety. Respondents reported purposeful routines such farming, exercise, and household responsibilities as effective coping strategies

> *“What is helping me now is I am doing farming these days…. it keeps me busy that I don’t have free time to spend, and also, I come back from the garden when am tired and have a lot of work at home like preparing food and other things.” (SU#26)*
>
> *“I go running in the morning and in the evening so I forget about everything that was going on, like if you were thinking about going and taking opium you will not. you just continue doing your own things and keeping yourself busy.” (SU#15)*

### Sub-theme 4.3: Community and family engagement for SUD recovery

Both HCWs and SUs highlighted family and community involvement as central to sustaining recovery. Participants proposed family counseling, community sensitization, and engagement of religious and local leaders to reduce stigma and enhance reintegration. SUs specifically proposed leveraging the influence of respected religious leaders such as pastors and imams to reframe SUD as a health condition rather than a moral failing.

> *“We need to conduct sensitization of the community about addictions, and this should start from schools.” (HCW#8)*

Respondents urged facilities to incorporate structured family counselling. They believed that when relatives understand addiction and receive guidance alongside the patient, they become allies in monitoring stressors and preventing relapse.

> *“…I feel the medical team should provide more education to the care takers of the patients who come to hospital to get treatment on the dangers of taking alcohol and its negative effects in a person’s life so that they can take back that message and also teach their children not to engage in alcohol taking.” (SU#9)*

### Sub-theme 4.4: Peer-led recovery; role of expert clients

In addition to formal treatment systems, both HCWs and SUs valued peers who had successfully recovered (“expert clients”) as credible mentors, viewed as relatable and trustworthy. Healthcare workers envisioned structured roles for them within the continuum of care, including training and empowering them to support the recovery of others.

> *“Since we’re complaining of human resource, we can use our very patients who are really complying with treatment to be the mentors in their communities… it gives them purpose and helps prevent relapse.” (HCW #10)*

### Sub-theme 4.5: Strengthening infrastructure and multidisciplinary care systems

HCWs emphasized the need to enhance facility infrastructure, such as erecting fences in some facilities, to prevent unauthorized access of addictive substances within treatment facilities. Additionally, participants highlighted the importance of enhancing technical capacity through staff training, recruitment and establishment of functional referral networks to deliver comprehensive multidisciplinary care addressing the root causes of addiction.

> *“That’s why on the referral system it should be both external referral from outside hospital from the community then also from within the hospital which is interdepartmental referral system for multidisciplinary approach.” (HCW#5)*

### Subtheme 4.6: Task sharing to strengthen community-based care continuity

In response to critical human resource shortages and fragmented follow-up systems, healthcare workers proposed task-sharing as a practical and cost-effective strategy to extend SUD care beyond hospital settings. They emphasized the pivotal role of CHWs often the first point of contact within the health system in supporting recovery, monitoring progress, and facilitating post-discharge follow-up. Participants suggested that CHWs and lower-level healthcare providers be trained and equipped to deliver basic psychosocial support, assist with rehabilitation, and strengthen relapse-prevention efforts at the community level.

> *“We often claim that we don’t have enough human resources or funding, but that’s not entirely true, we have people at the grassroots level, VHTs, get them to do trainings, motivate them, give them additional things, they can do this work very well.” (HCW #10)*

### Integrative thematic framework: linking barriers, vulnerabilities, and pathways to SUD recovery

The four meta-themes illustrated a self-reinforcing cycle shaped by systemic, clinical, social, and individual factors across the continuum of SUD treatment and recovery (Figure 1).

**Figure 1:**
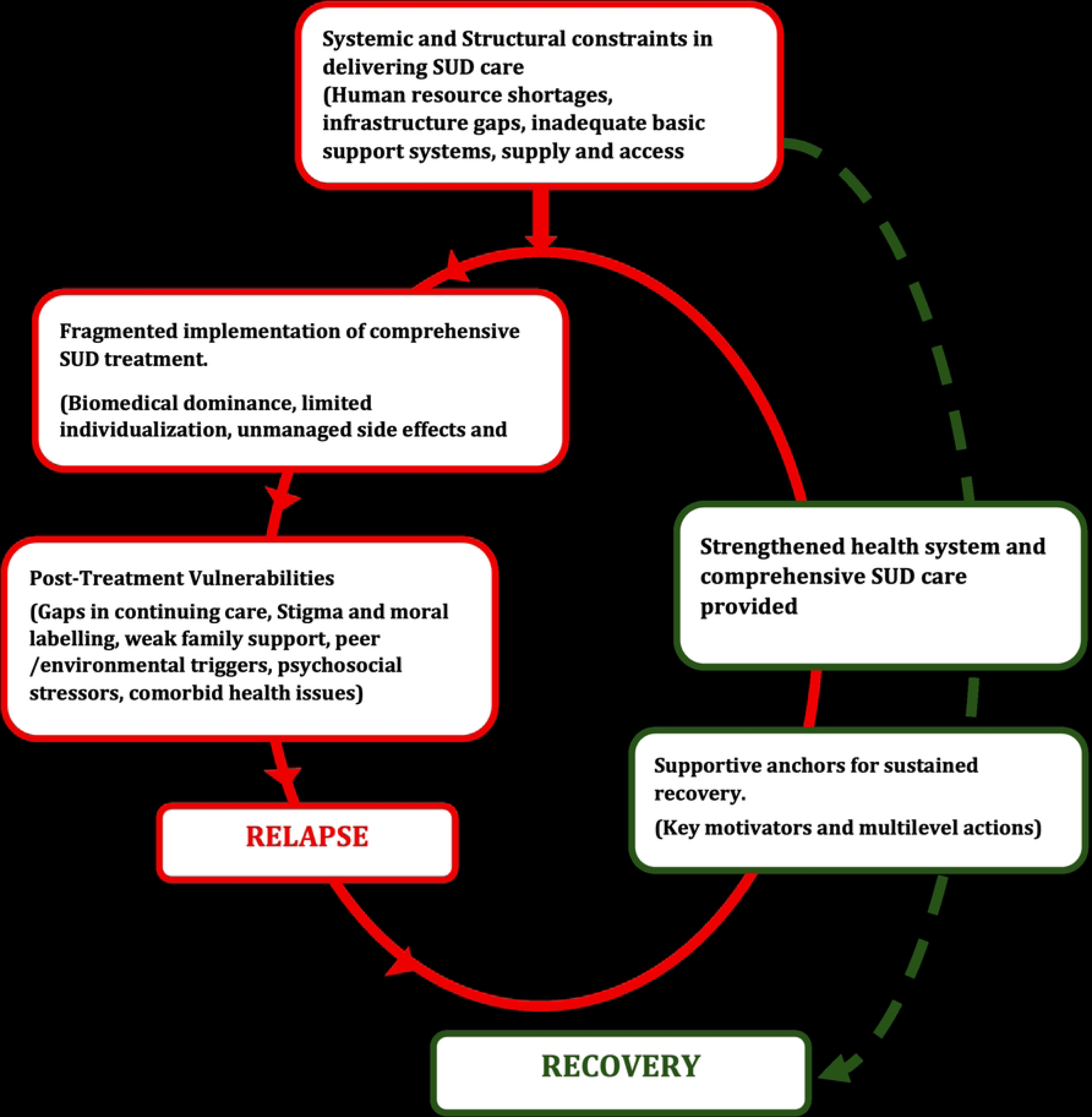
Self-reinforcing cycle of systemic and social dynamics of relapse and recovery in SUD care in Uganda

Systemic and structural limitations including inadequate human resources, infrastructural constrains, medication shortages, and geographic inequities undermined effectiveness and consistency of care. These weaknesses contributed to fragmented implementation of the biopsychosocial model, with biomedical approaches predominating while psychosocial support, individualized care, and long-term recovery planning were often neglected. Consequently, many patients were discharged with minimal or no structured aftercare, returning to unsupportive environments.

“Life Beyond the Ward” thus represented a fragile post-treatment context in which recovery was influenced by intersecting social, psychological, and structural conditions, such as persistent stigma and moral labelling, peer and environmental triggers, unstable family relationships, comorbid mental conditions, and limited access to follow-up or community-based care increasing the likelihood of relapse. Relapse in turn amplified demand on an already over-stretched system, further depleting the resources, and perpetuating the very gaps that precipitated relapse in the first place, creating a vicious loop. Relapse, while depicted linearly in the framework for conceptual clarity, is a multifactorial process shaped by these overlapping influences.

Nonetheless, both SUs and HCWs identified “pathways and supportive anchors for sustained recovery” including faith and spirituality, active lifestyle coping, peer-led initiatives, family and community engagement, and multidisciplinary care systems as critical enablers of resilience. Collectively, the meta-themes reveal how systemic and service-level barriers cascade into individual vulnerabilities, yet personal and social strengths can counterbalance these challenges to support sustained recovery.

## Discussion

This study explored the lived experiences of service users and health workers involved in substance use disorder treatment to explore multilevel barriers and facilitators shaping SUD treatment and recovery in Uganda. These findings add to limited African evidence on how health-system capacity, service delivery, and social context interact to influence SUD recovery.

Findings revealed a dynamic interaction between systemic, clinical, social, and individual factors that collectively shape recovery outcomes. The four meta-themes that were generated from this study illustrated a self-reinforcing cycle in which structural and resource constraints lead to fragmented SUD care that resolve acute crises but neglect relapse prevention strategies. Premature discharge of SUs into unsupportive environments and communities precipitated relapse which returned patients to an already over-stretched system, further depleting scarce resources. However, at every level of this cycle, participants also identified motivational and supportive anchors including spiritual and intrinsic motivation, family engagement, peer mentorship, and community engagement that could disrupt this cycle and shift the system towards sustained recovery.

One of the central findings of this study was the pervasive shortage of resources and infrastructure across SUD treatment facilities, which emerged as a key determinant of care quality and recovery outcomes. Both BNRMH and GRRH operated under significant human-resource constraints, coupled weak community-level linkages to support continuity of care. These deficits curtailed opportunities for individualized counseling, regular clinical reviews, and structured relapse-prevention planning which are core components of sustained recovery. The scarcity of trained personnel was compounded by frequent stockouts of psychotropic and anti-craving medications, inadequate therapeutic spaces, and overcrowded wards, collectively undermining therapeutic continuity and safety. Such resource deficiencies mirror broader patterns observed across sub-Saharan Africa, where limited health budgets, weak training pipelines, and fragmented mental health systems contribute to burnout, reduced treatment engagement, and high relapse rates (21,43–48). Nationally, readiness assessments have shown that only one third of facilities meet the minimum criteria to screen or manage SUDs, and fewer than 15% have staff trained in SUD diagnosis and management within the past two years (31).

Uganda’s mental health workforce density of 0.08 psychiatrists, 0.01 psychologists, 0.01 social workers, 0.01 occupational therapists, and 0.20 psychiatric clinical officers per 100,000 population (51) remains far below international standards. Globally, median numbers were 1.7 psychiatrists, 1.4 psychologists, 3.8 mental health nurses, and 0.7 social workers per 100,000 population(49). Collectively, these findings underscore that structural and workforce deficits, more than clinical protocols, shape treatment quality and continuity of care. Strengthening Uganda’s mental-health system will therefore require strategic investment in human resources, infrastructure, and supply-chain management to close the widening treatment gap for substance-use disorders.

To address critical workforce shortages and fragmented follow-up systems, HCWs recommended task-sharing with CHWs and other non-specialist providers. Global mental health literature and international guidelines endorse task-sharing as a practical and scalable strategy for LMICs (50,51). Evidence from successful interventions such as the Friendship Bench problem-solving therapy in Zimbabwe demonstrates that trained lay health workers can deliver effective psychological care with high retention and significant symptom reduction when supported by structured supervision and stakeholder engagement (52). In Uganda’s SUD treatment settings, where specialist staff are scarce and continuity of care remains weak, empowering CHWs and lower-level providers could extend care beyond hospital settings, enhance rehabilitation, and strengthen relapse-prevention efforts at the community level. However, as Le et al. (2022) noted, effective task-sharing requires addressing multilevel barriers including supervision, training, and referral coordination to ensure sustainability(51).

Uganda’s previous experience with CHW-led HIV and mental-health programs supports the feasibility of this model(53). Furthermore, a recent reviews found that non-specialist-delivered SUD interventions incorporating motivational interviewing, psychoeducation, and problem-solving techniques were effective across LMICs(54,55). Participants also highlighted the role of ‘expert client’ peers in long-term recovery who mentor others and bridge clinical and lived experience. Peer-facilitated recovery groups have demonstrated acceptability and improved functional outcomes suggesting their potential applicability for SUD care (56–58). Integrating structured peer-support systems within Uganda’s SUD care continuum could strengthen continuity between inpatient and community care, increase recovery capital, and promote more sustainable, patient-centered outcomes.

Medication shortages across facilities reflected systemic supply-chain gaps that undermine the continuity and quality of care for individuals with SUDs. Limited availability of essential psychotropic and anti-craving medications such as naltrexone and buprenorphine which was reported mirrors a broader pattern observed across LMICs, where frequent stockouts, weak logistics systems, and constrained budgets disrupt the management of chronic mental health conditions(49,59–61). In the absence of medicines, clinicians relied on psychosocial approaches, which, though adaptive, limited treatment effectiveness.(62). Medication unavailability contributes to disengagement from care, perceptions of ineffectiveness, and higher relapse rates among patients with substance-use and mental. The practice of asking patients to purchase medications privately though intended to bridge supply gaps, increased inequities by excluding those who could not afford from evidence-based treatments. Addressing these deficits requires strengthening procurement and supply-chain management systems for mental health and addiction medicines, integrating them into Uganda’s Essential Medicines List, and ensuring consistent procurement planning.

Although the biopsychosocial model was endorsed in practice, both HCWs and SUs described SUD care as primarily pharmacological, with counseling, occupational therapy, and family involvement inconsistently applied due to staffing shortages, time constraints, and limited psychosocial expertise. This biomedical dominance, while effective in stabilizing acute symptoms, neglects underlying behavioral and social determinants of substance use, resulting in transient recovery and high relapse risk (60). Similar patterns have been reported in other LMICs, where biomedical dominance persists(63,64), and rarely progresses to psychosocial or community rehabilitation despite policy promoting holistic care (65–67). Closely linked to this imbalance was a lack of individualized and participatory treatment planning, where standardized “one-size-fits-all” approaches replaced person-centered assessment and collaborative goal setting. Such limited personalization fails to capture the heterogeneity of substance-use trajectories, particularly among patients with co-occurring mental health conditions or severe social stressors, and has been associated with poor adherence and premature disengagement from care(21,68–70). Evidence shows that person-centred and collaborative models improve adherence, therapeutic alliance, and patient-reported recovery outcomes (71) whereas prescriptive or coercive practices, where patients are medicated without explanation and consent increased mistrust and reduced engagement(72). Additionally, sub-optimal in-patient treatment durations coupled with inadequate follow-up/continuing care post-discharge as was reported in other places in SSA (21) limit the effectiveness of interventions in addressing underlying issues and supporting long-term recovery (73–76).

Family support emerged as both a facilitator and barrier to recovery. Participants described families as critical sources of emotional stability, practical assistance, and motivation during treatment, findings consistent with prior research demonstrating that family engagement enhances treatment adherence and long-term recovery outcomes(77–80). However, for many, families were either absent, strained, or emotionally exhausted by repeated relapses and financial burdens of care. Family-related challenges, including stigma, misunderstanding of addiction, and limited involvement in treatment, have been linked to poor recovery outcomes in similar contexts (81,82). These findings align with previous studies indicating that families often lack accurate knowledge of substance-use disorders, which limits their capacity to provide consistent support (83,84).

To strengthen recovery systems, families should be viewed not as part of the problem but as integral partners in treatment and aftercare. Evidence indicates that family-inclusive interventions produce greater improvements in abstinence and functioning compared to individual counseling alone, leveraging relational support to sustain behavior change(85–88). Integrating structured family psychoeducation, counselling, and communication skills into routine SUD care consistent with WHO mhGAP guidance could enhance understanding, reduce stigma, and foster a collaborative recovery environment(89).

Moral stigma, stigmatizing language and negative stereotypes perpetuated by both HCWs and the broader community contributed significantly to social isolation and negative treatment perceptions. Labeling individuals as "addicts" or "abusers" not only undermines recovery efforts but also perpetuates self-stigma, which can deter them from accepting treatment (90–93). The fear of being labeled or judged negatively by society leads to secrecy and reluctance to disclose struggles with SUD and seek treatment. This reluctance stems from concerns about potential repercussions, including social exclusion, discrimination, and loss of respect. (94).

A notable finding from this study was the economic entanglement between substance use and livelihood, particularly in rural settings where alcohol brewing or vending provided a primary or sole income source for some participants. Similar patterns have been reported in other African contexts, where the informal alcohol economy sustains entire households despite its health and social harms (95,96). For individuals whose livelihoods depend on production or sale of intoxicating substances, abstinence becomes nearly impossible, as continued use is reinforced by economic necessity and social normalization within these networks. The entanglement of substance use with livelihood and survival strategies underscores the need for integrated approaches that combine economic empowerment with harm-reduction interventions, addressing both the social determinants of addiction and the structural barriers to recovery. Evidence from Kenya demonstrates that livelihood-replacement programs coupled with motivational interviewing can reduce hazardous drinking and enhance social reintegration among participants (95,97). These findings highlight that treatment strategies focusing solely on clinical stabilization, without addressing underlying economic vulnerability, risk short-term success followed by relapse once individuals return to precarious living conditions.

Both SUs and HCWs described peer networks as powerful drivers of relapse, consistent with the social-learning theory, which explains that behavior is shaped and maintained through observing others and receiving reinforcement from significant peers(98). Some studies have indicated that social interactions, particularly with peers who engage in substance use, can exacerbate the risk of relapse following treatment. This influence is multifaceted, involving both direct peer behaviors and the broader social environment (99–101). Integrating peer-refusal skills training with routine counselling has been reported to enhance positive recovery outcomes(102,103). Conversely, the importance of faith-based coping to prevent relapse as indicated by participants is consistent with other longitudinal Ugandan data linking religiosity to reduced harmful alcohol use(104,105). Mechanistically, spirituality contributed to personal recovery capital which included self-efficacy, meaning-making and social capital where spiritual leaders provided the accountability networks.

The intersection between poverty, unemployment, and substance use also emerged as a recurring stressor. Many participants described idleness and economic insecurity as triggers for relapse, consistent with prior research linking social exclusion and relapse vulnerability in LMIC contexts. Respondents who mentioned that they replaced idle time with farming, running or church duties described reduced craving for substances. These narratives align with behavioural-activation theory which emphasizes that engaging in positive, value-driven activities, such as physical activity can serve as a beneficial adjunct to traditional treatment methods. A randomized controlled trial by Daughters et.al., (2018) demonstrated that participants in a behavioral activation treatment had significantly higher likelihood of abstinence and reduced adverse consequences from substance use(106). Additionally, a systematic review indicated that approximately 75% of studies found a decrease in substance use following physical activity interventions, highlighting its effectiveness as a complementary therapy (107).

The figure developed from this study (Figure 1) illustrated how systemic constraints and social stressors form a vicious cycle of relapse and resource depletion. However, it also showed how multi-level motivators including spiritual, familial, peer-led mentorship, community engagement, and multisectoral system-strengthening initiatives can break into this cycle, transforming it into a resilient, recovery-oriented system.

While this study provides valuable insights into the multilevel barriers and facilitators shaping treatment journeys through HCWs and SUs perspectives, findings should be interpreted with acknowledgement of some limitations. The limited number of participants purposively selected could consequently constrain transferability and generalizability of findings to other geographical settings or primary-care levels. Factors unique to these settings may influence the identified themes, limiting their applicability to other contexts. Also, data were generated through single, cross-sectional interviews, precluding observation of how perceptions could evolve over time.

The dual role of some clinicians who were research assistants in the initial cohort study could have introduced social-desirability bias, particularly regarding HCW accounts of service quality since residual bias cannot be excluded. The perspectives shared by HCWs may be influenced by their specific roles within the healthcare system and their personal experiences, which could introduce bias into the data.

## Conclusions

This study highlights how structural, clinical, and social factors intersect to shape substance SUD treatment and recovery in Uganda. Systemic resource shortages, workforce gaps, and fragmented psychosocial support undermine the implementation of the biopsychosocial model, resulting in care that resolves acute crises but rarely sustains recovery. Medication shortages, brief inpatient stays, and limited community reintegration further perpetuate relapse and cyclical readmissions. Yet within these constraints, participants identified protective anchors such as spiritual faith, peer mentorship, family involvement, and community engagement that can be strengthened to promote recovery and resilience.

These findings emphasize the need to shift towards a continuum-of-care approach that integrates facility-based and community-level interventions. Policy priorities should include expanding human resource capacity through task-sharing, embedding psychosocial and family-inclusive interventions within standard care, and ensuring reliable access to essential medications. Addressing social determinants such as poverty, stigma, and economic dependency on substance-related livelihoods will also be critical to sustaining recovery beyond treatment settings.

## Data Availability

The data underlying this study contain sensitive personal and clinical information from human participants and cannot be shared publicly due to ethical restrictions. De-identified data may be made available to qualified researchers upon request. Requests for access should be directed to the corresponding author at.

## Acknowledgments

The authors would like to extend their sincere appreciation to the Makerere University School of Public Health for the financial support provided through the seed grants programme (MakSPH-GRCB/2022/01), which made this study possible. We also wish to thank Wim Demey for his valuable contributions and support throughout this work. Our gratitude goes to the study participants for their time and willingness to share their experiences, as well as to the dedicated research assistants whose commitment and hard work were essential to the successful completion of the study.

